# Tumor Mutational Burden Predicts Survival In Patients With Low Grade Gliomas Expressing Mutated IDH1

**DOI:** 10.1101/2020.01.20.20016766

**Authors:** Mahmoud S Alghamri, Rohit Thalla, Ruthvik Avvari, Ali Dabaja, Ayman Taher, Lili Zhao, Maria G Castro, Pedro R Lowenstein

## Abstract

Gliomas are the most common primary brain tumors. High Grade Gliomas have a median survival of 18 months, while Low Grade Gliomas (LGG) have a median survival of ∼7.3 years. Seventy-six percent of patients with LGG express mutated isocitrate dehydrogenase (mIDH1) enzyme (IDH1^R132H^). Survival of these patients ranges from 1-15 years, and tumor mutational burden ranges from 8 to 447 total somatic mutations per tumor. We tested the hypothesis that the tumor mutational burden would predict survival of patients with tumors bearing mIDH1^R132H^. We analyzed the effect of tumor mutational burden on patients’ survival using clinical and genomic data of 1199 glioma patients from The Cancer Genome Atlas and validated our results using the Chinese Glioma Genome Atlas. High tumor mutational burden negatively correlates with survival of patients with LGG harboring IDH1^R132H^ (p<0.0001). This effect was significant for both Oligodendroglioma and Astrocytoma LGG-**m**IDH1 patients. In the TCGA data, median survival of the high mutational burden group was 76 months, while in the low mutational burden group it was 136 months; p<0.0001. There was no differential representation of frequently mutated genes (e.g., *TP53, ATRX, CIC, FUBP*) in either group. Gene set enrichment analysis revealed an enrichment in Gene Ontologies related to Cell cycle, DNA damage response in high vs low tumor mutational burden. Finally, we identified a 19 gene signature that predicts survival for patients from both databases. In summary, we demonstrate that tumor mutational burden is a powerful, robust, and clinically relevant prognostic factor of median survival in mIDH1 patients.

## INTRODUCTION

Low grade gliomas (LGGs) are slow-growing brain tumors that occur in early adult life and can progress to high grade gliomas (1). Molecular characterizations coupled with the histological classification revealed that a mutation in isocitrate dehydrogenase (mIDH), IDH1^R132H^, is the main genetic lesion in LGG patients (2-5). Somatic mutation in IDH1, and far less common IDH2, results in excessive production of 2 hydroxyglutarate (2HG) (6-8). 2HG is a potent and competitive inhibitor of α-ketoglutarate (αKG) dependent dioxygenases, which responsible for demethylation of DNA and histones (4, 9). The ensuing hypermethylation phenotype triggers epigenetic reprogramming of the glioma cells’ transcriptome (3, 10-13). The consequences of mIDH1 in glioma cells contribute to cancer development and progression not only by disrupting cell metabolism but also by altering the epigenetic landscape. Metabolically, IDH1 is one of the enzymes that encodes an irreversible reaction in the Tricyclic acid (TCA) cycle. Disruption the IDH1 reaction results in defective mitochondrial oxidative phosphorylation, glutamine metabolism, lipogenesis, glucose sensing, and altering of cellular redox status (8, 14-18). IDH1 also inhibits glioma stem cell differentiation (4, 8), upregulates vascular endothelial growth factor (VEGF) to promote tumor microenvironment formation (19, 20), and produces high levels of hypoxia-inducible factor-1α (HIF-1α) to promote glioma invasion (18, 21, 22), which ultimately leads to glioma progression. We have acquired a tremendous insight of the molecular mechanisms of genetic alterations associated with malignant transformation of IDH mutations. This includes activation of NOTCH1, RTK-RAS-PI3K, and Myc-RB1 signaling and/or deleted region of the CDKN2A/2B locus, all of which were found to be altered in progressed mIDH1 GBM samples compare to their lower grade counterparts (2, 23-25). Nevertheless, the exact mechanism that impacts mIDH1 patient survival is still under debate.

In this regard, a significant advance in recent years has identified a set of genetic lesions that are characteristic of mIDH1 and correlate with clinical outcome. This was done in hopes for better prediction of tumor behavior and outcome, including identification of secondary mutations, genetic alterations, methylation patterns, and multivariate prognostic models (26-29). Within the group of IDH-mutant gliomas, presence of 1p/19q co-deletion (IDH-mutant–codel glioma) may present an additional prognostic marker separate from IDH-mutant glioma with intact 1p/19q chromosome arms (IDH-mutant–non-codel glioma). Other genetic alteration affect IDH1 glioma survival includes mutation of *PIK3CA* and *PIK3R1*, and deletion of *CDKN2A* in Astrocytoma mIDH1 (30, 31).

Given the high level of inter-tumor heterogeneity inferred from the presence of variety of genetic lesions and the fact that the IDH1 mutant tumors are resistant to radiotherapy than the wtIDH1 (32, 33), it is tempting to speculate that tumor mutational burden may be a strong predictor for mIDH1 patient prognosis. In most solid cancers, the mutational load negatively impacts the patient survival (34) and it is used as indicator of rapid tumor progression.

In the present study, we analyzed the publicly available TCGA and CGGA datasets in order to correlate the tumor mutational burden with overall survival in mIDH1 LGG subtypes. We found that increasing tumor mutational burden negatively impacts survival of LGG but not GBM. Further analysis of LGG patients revealed that the effect of tumor mutational burden on survival is unique for patients with mIDH1 but not wtIDH1. Moreover, based on the prognosis, we constructed a high-risk gene set that predict poor prognosis in patients with IDH mutation. These data suggest that tumor mutational burden is an independent prognostic factor for glioma patients with IDH1 mutation and can be used as a predictor of patient survival.

## RESULTS

### Tumor mutational burden unexpectedly predicts increased aggressive clinical course only in LGG-mIDH1

We analyzed the clinical data of 1199 patients form TCGA. Among the TCGA patients we used data from 799 patients for whom there are available complete mutational data. All patients’ clinical and mutational data are summarized in Table1 and Table2. We asked if the tumor mutational burden impacts glioma patients’ survival. We analyzed the tumor mutational burden in high grade glioma (HGG) and low grade glioma (LGG) in TCGA and validated our results utilizing the CGGA database. We classified all patients according to the tumor mutational burden into two groups (high vs low), based on the median number of mutations for all patients in a group. We found that overall mutational load was higher in HGG compare to the LGG, regardless of the IDH1 mutation (supplement 1A, B). We then applied the ‘life test’ procedure to compare the survival probabilities between patients with high vs low tumor mutational burden. Surprisingly, there was no differences in survival between HGG with high tumor mutational burden as compared to HGG with low tumor mutational burden (Fig 1A).

**Table 1.** Clinical Characteristic of TCGA patients

|  |  | **LGG** |  |  |  | **GBM** |  |
| --- | --- | --- | --- | --- | --- | --- | --- |
| **Descriptors** |  | wtIDH1 High | wtIDH1 Low | mIDH1 High | mIDH1 Low | wtIDH1 High | wtIDH1 Low |
| **Gender, n(%)** | Male | 30 (51.72) | 34 (59.65) | 121 (60.5) | 100 (50) | 79 (56.43) | 100 (71.43) |
|  | Female | 28 (48.28) | 23 (40.35) | 79 (39.5) | 100 (50) | 61 (43.57) | 40 (28.57) |
| **Age, median (25<sup>th</sup>-75<sup>th</sup>)** |  | 58 (49-63) | 41 (29-57) | 44 (37-55) | 34 (29-41) | 58 (50-67) | 65 (58-73) |
| **Radiation Therapy, n(%)** | Yes | 45 (77.59) | 30 (52.63) | 123 (61.5) | 98 (49) | 106 (75.71) | 111 (79.29) |
|  | No | 8 (13.79) | 21 (36.84) | 67 (33.5) | 90 (45) | 22 (15.71) | 24 (17.14) |
|  | NA | 5 (8.62) | 6 (10.53) | 10 (5) | 12 (6) | 12 (8.57) | 5 (3.57) |
| **TMZ, n(%)** |  | 21 (36) | 25 (43) | 52 (26) | 42 (21) | 41 (29) | 44 (31) |
| **Race, n(%)** | Asian | 1 (1.72) | 1 (1.75) | 5 (2.5) | 1 (0.5) | 2 (1.43) | 3 (2.14) |
| **Race, n(%)** | Black or African American | 5 (8.62) | 4 (7.02) | 7 (3.5) | 5 (2.5) | 10 (7.14) | 7 (5) |
|  | White | 52 (89.66) | 49 (85.96) | 187 (93.5) | 187 (93.5) | 126 (90) | 126 (90) |
|  | Other | 0 (0) | 3 (5.26) | 1 (0.05) | 7 (3.5) | 2 (1.43) | 4 (2.86) |
| **Median Survival (days)** |  | 686 | 1578 | 2282 | 4084 | 427 | 414 |
| **Histology (LGG), n(%)** | Astrocytoma | 32 (55.17) | 26 (45.61) | 67 (33.5) | 69 (34.5) | NA | NA |
|  | Oligoastrocytoma | 12 (20.69) | 9 (15.79) | 50 (25) | 59 (29.5) | NA | NA |
|  | Oligodendroglioma | 14 (24.14) | 22 (38.6) | 83 (41.5) | 72 (36) | NA | NA |
| **Histology (GBM), n(%)** | Glioblastoma Multiforme (GBM) | NA | NA | NA | NA | 8 (5.71) | 3 (2.14) |
|  | Treated Primary GBM | NA | NA | NA | NA | 1 (0.71) | 3 (2.14) |
|  | Untreated Primary (De Novo) GBM | NA | NA | NA | NA | 131 (93.57) | 134 (95.71) |
| **1P 19Q n(%)** |  | 3 (5.17) | 13 (22.81) | 78 (39.00) | 77 (38.5) | 0 (0) | 0 (0) |

LGG: Low grade glioma, mIDH: mutation in isocitrate dehydrogenase. TMZ: Temazolamide

**Table 2.** Clinical Characteristic of CGGA patients

|  |  | **LGG** |  |  |  | **GBM** |  |
| --- | --- | --- | --- | --- | --- | --- | --- |
| **Descriptors** |  | wtIDH1<br>High | wtIDH1<br>Low | mIDH1<br>High | mIDH1<br>Low | wtIDH1<br>High | wtIDH1<br>Low |
| **Gender, n(%)** | Male | 17 (47.22) | 22 (61.11) | 30 (53.57) | 44 (78.57) | 22 (55.00) | 20 (50.00) |
|  | Female | 19 (42.88) | 14 (38.99) | 26 (46.43) | 12 (21.43) | 18 (45.00) | 20 (28.57) |
| **Age at Diagnosis, median (25th and 75th percentile)** |  | 40 (34-48) | 38 (28-43) | 42 (35-45) | 36 (31-42) | 56 (45-63) | 49 (34-60) |
| **Radiation Therapy, n(%)** | Yes | 27 (75) | 27 (75) | 37 (66.07) | 41 (73.21) | 28 (63.64) | 31 (77.5) |
|  | No | 5 (13.89) | 5 (13.89) | 8 (14.29) | 8 (14.29) | 3 (6.82) | 4 (10) |
|  | NA | 4 (11.11) | 4 (11.1) | 11 (19.64) | 7 (12.5) | 9 (20.45) | 5 (12.5) |
| **Median Survival (days)** |  | 1342 | 2210 | 2982 | Undefined | 310 | 765 |
| **Histological Type, n(%)** | Astrocytoma | 12 (33.33) | 16 (44.44) | 17 (30.36) | 7 (12.5) | NA | NA |
|  | Oligoastrocytoma | 20 (55.56) | 17 (47.22) | 29 (51.79) | 33 (58.93) | NA | NA |
|  | Oligodendroglioma | 4 (11.11) | 3 (8.33) | 10 (17.86) | 16 (28.57) | NA | NA |
| **Subtype, n(%)** | Glioblastoma Multiforme (GBM) | NA | NA | NA | NA | 24 (60) | 20 (50) |
|  | Primary GBM | NA | NA | NA | NA | 16 (40) | 20 (50) |
|  | Primary | 23 (63.89) | 25 (69.44) | 27 (48.21) | 51 (91.07) | 16 (40) | 20 (50) |
|  | Recurrent | 13 (36.11) | 11 (30.56) | 29 (51.79) | 5 (8.93) | 24 (60) | 20 (50) |

LGG: Low grade glioma, mIDH1: mutation in isocitrate dehydrogenase.

**Figure 1.**
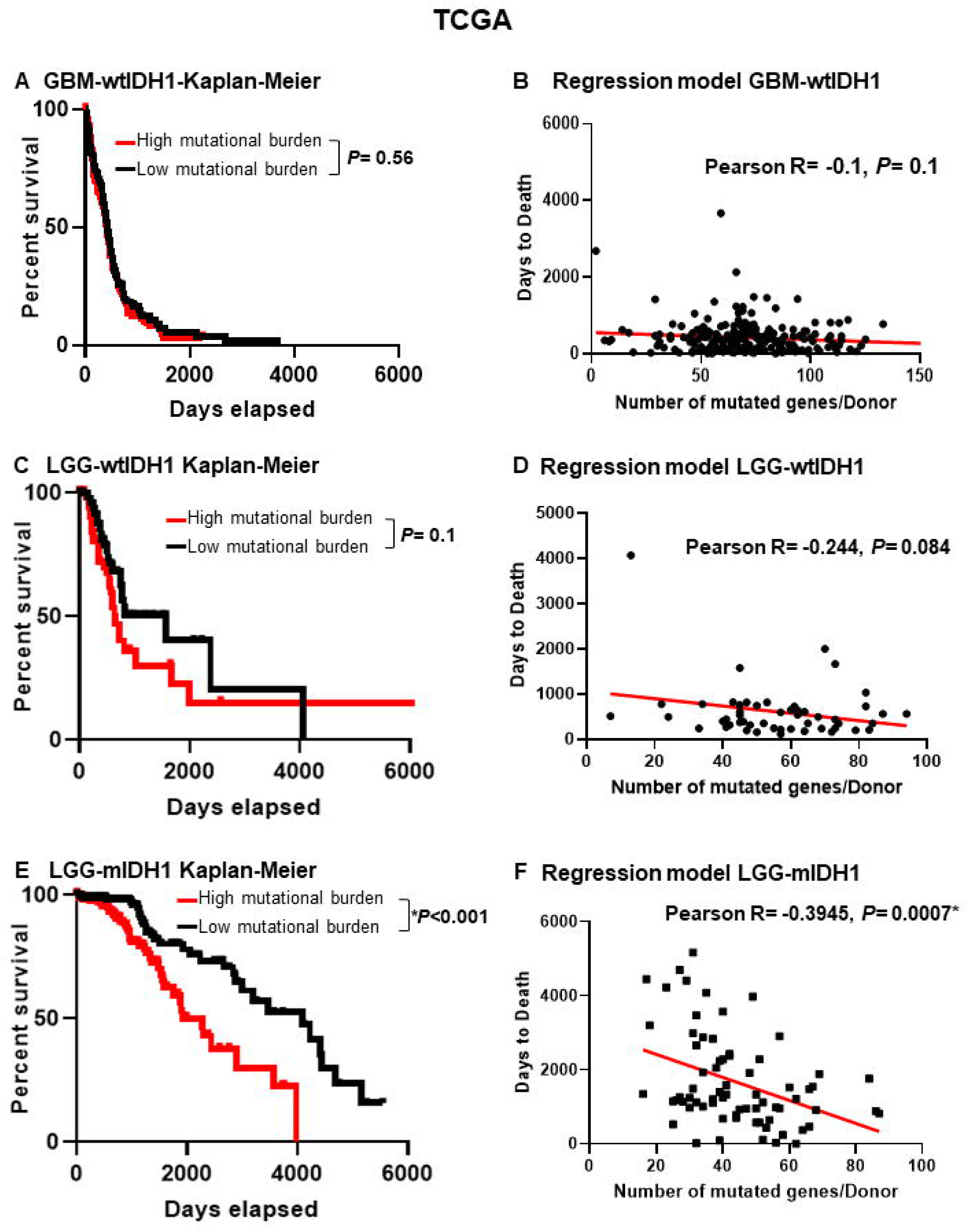
Poor prognosis in mIDH1 patients from TCGA with high mutational burden. **(A)** Kaplan-Meier survival of GBM-**wt**IDH1 patients with high mutational burden (red) or low mutational burden (black) from TCGA. There was no significant difference in survival between groups (Hazard Ratio (HR)=0.925, CI [0.651-1.313]) **(B)** Linear regression model of patient’s “days to death” vs mutational burden/patient in GBM-wtIDH1. There was no correlation between patients’ “days to death” and mutational load. **(C)** Kaplan-Meier curves of LGG-**wt**IDH1 patients classified according to mutational burden and IDH1 mutation in TCGA. There is no difference in survival between LGG-**wt**IDH1^**high**^ and LGG-**wt**IDH1^**low**^ (HR=0.686, CI [0.393-1.196]) (**D)** Linear regression model of patient’s “days to death” vs mutation burden/patient in LGG-wtIDH1. There was no correlation between patients’ “days to death” and mutational load. **(E)** LGG-**m**IDH1^**high**^ have statistically significantly decreased median survival as compared to LGG-**m**IDH1^**low**^ (HR=0.486, CI [0.294-0.794]). **(F)** Linear regression model of patient’s “days to death” vs mutation burden/patient in LGG-mIDH1. There is a significant correlation between patients’ days to death and mutation load/patient in LGG-mIDH1 only (R=-0.39, P<0.001).

In contrast, high tumor mutational burden significantly decreased the MS of LGG patients (supplement 2A). We stratified the LGG patients in terms of the IDH1 mutation and found no significant differences in MS between patients with tumors expressing wild type IDH1 and high number of mutations (LGG-**wt**IDH1^**high**^) as compared to patients with LGG-**wt**IDH1^**low**^ tumors (Fig 1C). Since the majority of LGG patients harbor a mutation in IDH1, that provides a strong survival benefit, we tested if the effect of a high tumor mutational burden on survival can be detected in LGG patients whose tumors harbor the IDH1 mutation. In the LGG-**m**IDH1 group, high tumor mutational burden was negatively correlated with patients’ prognosis (Fig 1E). In HGG patients with **m**IDH1 tumors, tumor mutational burden was also negatively correlated with survival, even though the number of patients available to be studied was low (n=14) (supplement 2A).

Within the group of LGG patients, oligodendrogliomas (Oligo) have a better overall survival (OS) than astrocytomas (Astro). We therefore determined if the tumor mutational burden had an effect on survival in each LGG subtype. Tumor mutational burden negatively impacted the prognosis of both Oligo and Astro glioma patients (supplement 2C). There was also no correlation with patient age, gender, treatment or histological classification of LGG-**m**IDH1^**high**^ vs LGG-**m**IDH1^**low**^ on OS (Table 3). We validated these results by analyzing the role of tumor mutational burden through the web-based analytical tool, i.e., the *cBioPortal* database (35, 36). The results obtained confirm our analysis (supplement 3A-C).

**Table 3.** Univariate Cox analysis in TCGA LGG-mIDH1

|  | **LGG-mIDH1<sup>low</sup>** |  |  | **LGG-mIDH1<sup>high</sup>** |  |  |
| --- | --- | --- | --- | --- | --- | --- |
|  | Mean | Upper | Lower | Mean | Upper | Lower |
| **Gender** | 1.176 | 2.399 | 0.576 | 0.8065 | 1.501 | 0.4335 |
| **Age** | 0.7026 | 1.91 | 0.2585 | 0.8382 | 1.578 | 0.4452 |
| **Radiotherapy** | 1.655 | 3.569 | 0.7676 | 1.352 | 2.615 | 0.699 |
| **Oligodendro** | 0.5396 | 1.13 | 0.2576 | 1.285 | 2.391 | 0.6901 |
| **Oligoastro** | 1.26 | 2.86 | 0.5553 | 0.5971 | 1.108 | 0.2452 |
| **Astro** | 1.676 | 3.774 | 0.7444 | 1.387 | 2.683 | 0.7167 |
| **1P 19Q Co-deletion** | 0.7042 | 1.494 | 0.3320 | 0.8084 | 1.517 | 0.4307 |
| **TMZ** | 0.9604 | 2.078 | 0.4438 | 1.746 | 3.145 | 0.8928 |

LGG: Low grade glioma, Oigodendro: Oligodendroglioma, Astro: Astrocytoma, Oligoastro:
Oligoastrocytoma, mIDH1: mutation in isocitrate dehydrogenase. TMZ: Temazolamide

We further validated the effect of tumor mutational burden in LGG-**m**IDH1 using two approaches.

First, we tested if there was a correlation between the survival rate and tumor burden in all three tumor types. We applied a linear regression model to see if we could predict the “days to death” (dependent variable) of glioma patients based on the tumor mutational burden (independent variable). Since an endpoint is required for linear regression, we only considered the deceased patients in this analysis. Linear regression modeling showed a significant dependency of patients “days to death” on tumor mutational burden only in patients with LGG-**m**IDH1 tumors (Fig 1B, D, F).

We then validated our TCGA results by studying 286 patients from the CGGA database to further corroborate our hypothesis using a separate data set. In agreement with the results from the TCGA, no difference in survival was seen between GBM-**wt**IDH1^**high**^ and GBM-**wt**IDH1^**low**^ nor LGG-**wt**IDH1^**high**^ and LGG-**wt**IDH1^**low**^ (Fig 2A, B). In patients with LGG-**m**IDH1 tumors, however, LGG-**m**IDH1^**high**^ have significantly poorer prognosis compared to LGG-**m**IDH1^**low**^ (Figure 2C). These data confirm the effects of tumor mutational burden on survival of patient with LGG expressing the IDH1 mutation, in two international unrelated databases.

**Figure 2.**
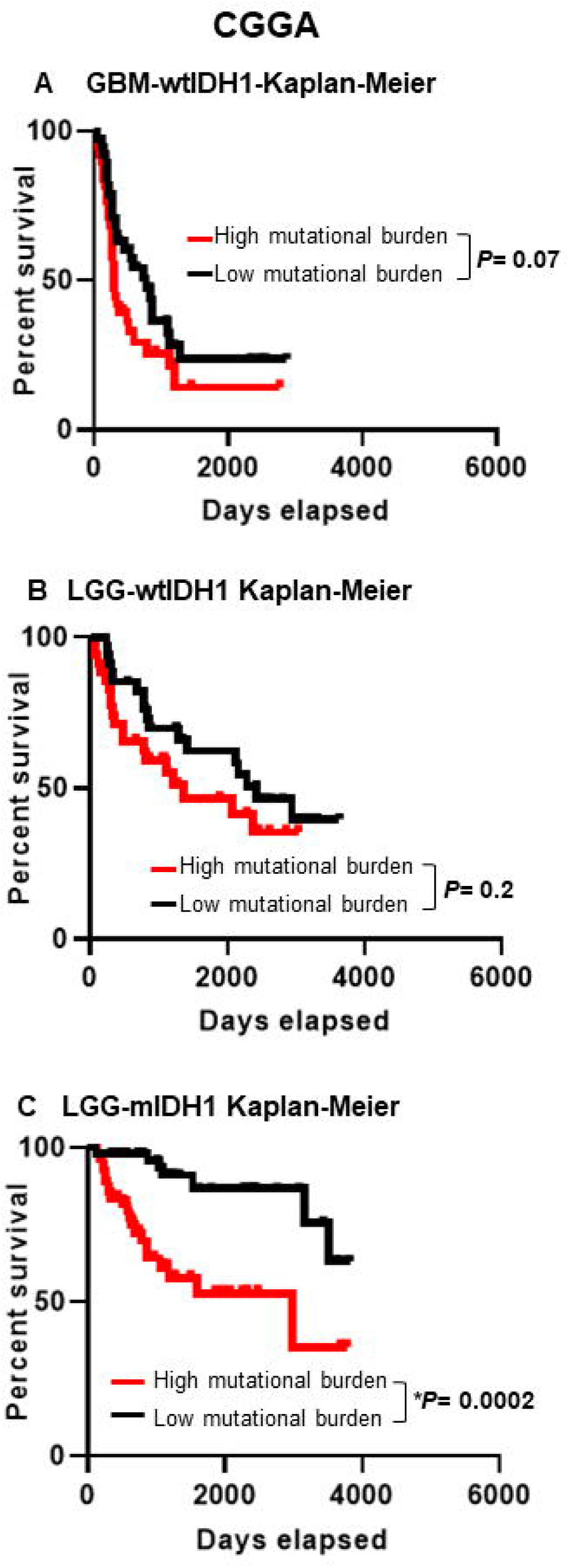
Poor prognosis in mIDH1 patients with high mutational burden in CGGA. **(A)** Kaplan-Meier survival of GBM-**wt**IDH1 patients with high mutational burden (red) or low mutational burden (black) in CGGA (HR=0.91, CI [0.651-1.18]). There is no statistically significant difference between both groups. **(B)** Kaplan-Meier of LGG-**wt**IDH1 patients classified according to mutational burden and IDH1 mutation in CGGA. There is no difference in survival between LGG-**wt**IDH1^**high**^ LGG-**wt**IDH1^**low**^ (HR=0.644, CI [0.329-1.18]). There is no statistically significant difference between both groups. **(C)** Kaplan-Meier of LGG-**m**IDH1 patients classified according to mutational burden and IDH1 mutation in CGGA. There is a statistically significant difference in survival between LGG-**m**IDH1^**high**^ vs LGG-**m**IDH1^**low**^ (HR=0.236, CI [0.11-0.5]) (*p*<0.001).

### Association of genetic alterations with overall survival (OS)

To determine if the differences in tumor mutational load could be explained by changes in the frequency of individual mutations, mutational signatures (COSMIC), or copy number variation, each of these was analyzed in detail. We found that nine out of the top ten most commonly mutated genes are present at the same frequency in both groups. These genes are (IDH1, TP53, ATRX, CIC, FUBP1, TTN, PIK3CA, MUC16, Notch1) (Fig 3A, B). None of these mutations impart a significant difference in OS between LGG-**m**IDH1^**high**^ and LGG-**m**IDH1^**low**^ (Fig 4A). The COSMIC database has categorized 30 reference mutation spectra signatures based on the analysis of 40 distinct types of human cancer (37). The genetic mutation signatures across all patients within the LGG-**m**IDH1^**high**^ and LGG-**m**IDH1^**low**^ groups were identical (Fig 3A, B, bottom plots). The most common signatures in either group were 1, 6, and 15 (Fig 3 A, B, supplement 4).

**Figure 3.**
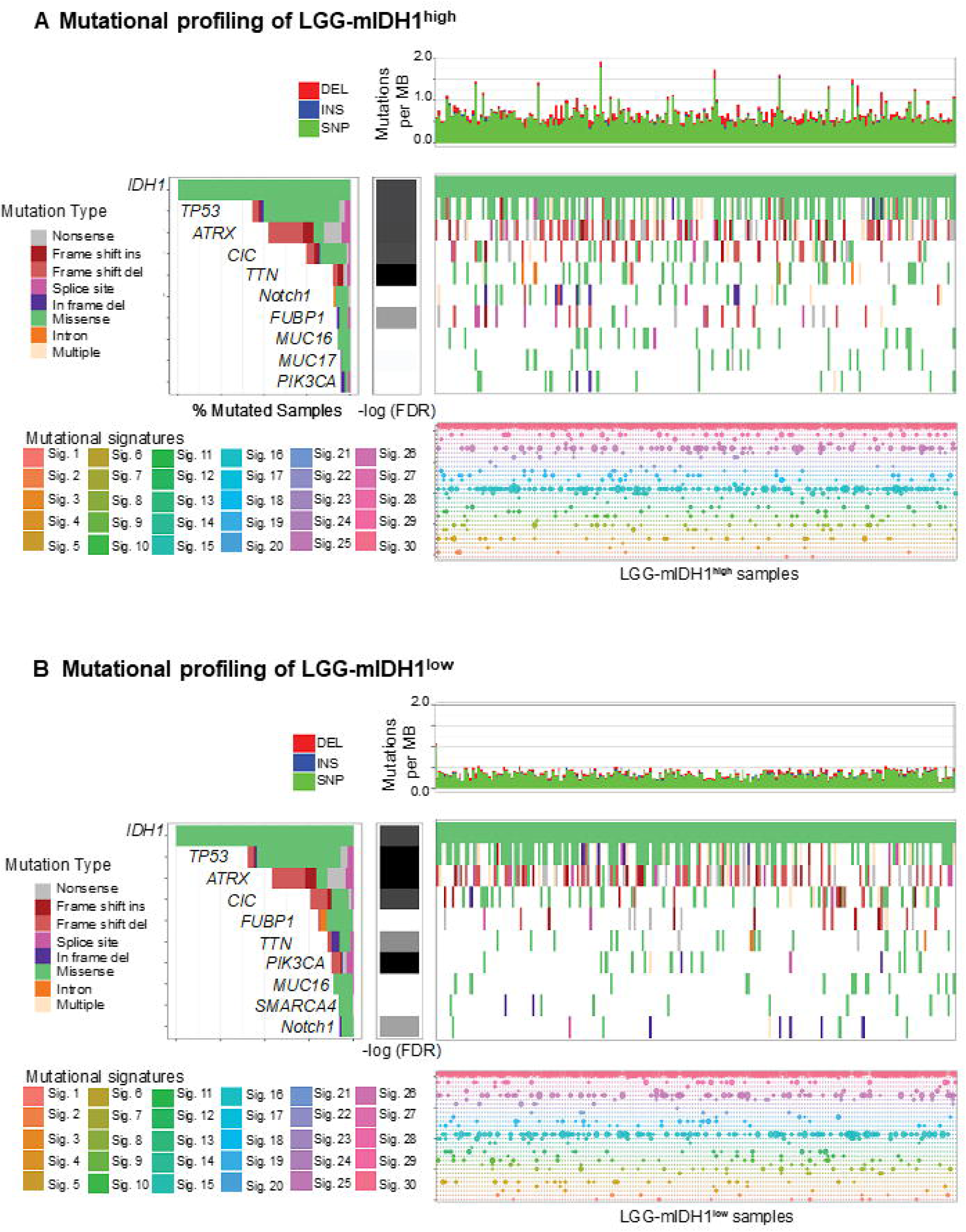
Somatic genetic alterations identified in LGG-mIDH1 according to the tumor mutational burden. The upper plot shows the mutation rate for each tumor sample. Middle plot: Heatmap of most frequent somatic mutations identified in LGG-**m**IDH1^**high**^ **(A)** vs LGG-**wt**IDH1^**low**^ **(B)**. Mutation types are color-coded according to the legend. Lower plot: mutational signature analysis between LGG-**m**IDH1^**high**^ vs LGG-**m**IDH1^low^, respectively.

**Figure 4.**
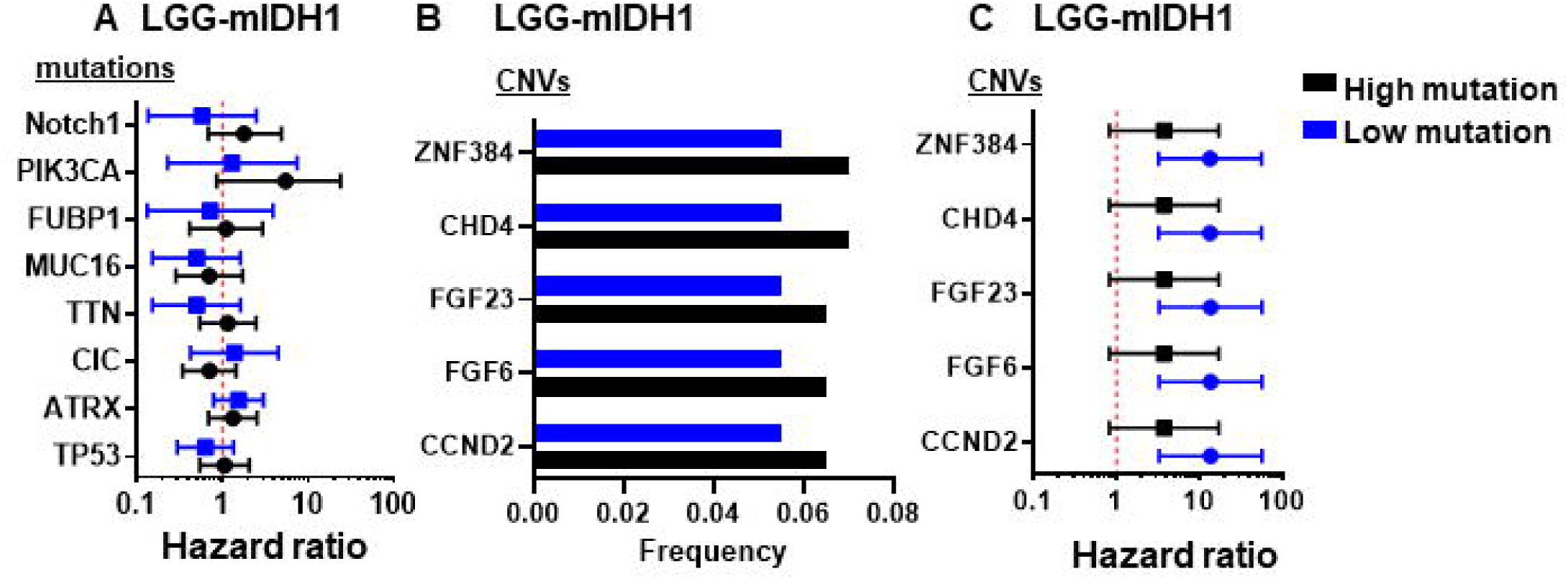
Mutations frequency and Hazard ratios (HRs) for OS in Cox regression model in LGG-mIDH1^high^ vs LGG-mIDH1^low^. **(A)** HRs for OS in Cox regression model according to the presence or absence of the frequently mutated genes among the groups. **(B)** The most frequent CNVs in high vs low mutation in LGG-**m**IDH1. **(C)** HRs for OS in Cox regression model according to the presence or absence of the frequently CNVs among the groups.

We then investigated the effect of the most frequent copy number variations (CNVs) on LGG-**m**IDH1^**high**^ and LGG-**m**IDH1^**low**^. In all LGG-**m**IDH1, the most frequent CNVs are found in the following genes: CCND2, FGF6, FGF23, CHD4, ZNF384 (Fig 4B). All of these CNVs have a negative impact on the OS of LGG-**m**IDH1^**low**^ (Figure 4B, C). However, these CNVs are all present in a small group of patients (5.5% of LGG-**m**IDH1^**low**^, and 6.5% of LGG-**wt**IDH1^**high**^). Overall, these data suggest that tumor mutational burden is a unique and independent prognostic factor that negatively impacts LGG-**m**IDH1 patient survival.

### Unique gene ontology (GO) groups are enriched in the LGG-mIDH1^high^

Since the hyper-mutational phenotype impacts LGG-**m**IDH1 but not the LGG-**wt**IDH1, we hypothesized that LGG-**m**IDH1^**high**^ is associated with a differential gene expression profile, which could be used to predict patients’ outcome. We performed the differential gene analysis based on tumors with either high or low tumor mutational burden. Using an FDR of 0.01 as the lower limit of significance we identified 1585 genes that were upregulated, and 1865 genes that were downregulated in the LGG-**m**IDH1^**high**^. We then performed gene set enrichment analysis (GSEA) to evaluate the functional aspects of the differentially expressed genes. We compared the differences between significant GOs of LGG-**wt**IDH1^**high vs low**^ and LGG-**m**IDH1^**high vs low**^. Results suggested that high tumor mutational burden was associated with upregulation of DNA repair, cell cycle-related processes, and chromosomal remodeling in LGG-**wt**IDH1^**high vs low**^ (Fig 5A, C, supplement 5). In LGG-**m**IDH1^**high vs low**^, there was enrichment in GO families belonging to regulation of cell cycle processes and DNA mismatch repair pathways (Fig 5D, supplement 6). Interestingly, there was a positive enrichment of GO families that belong to RNA and pre-RNA processing in LGG-**m**IDH1^**high vs low**^ (Fig 5D). This is consistent with recent report highlighting the effect of enrichment in RNA processing associated GOs on LGG patients’ survival (38). We further validated the GSEA analysis using LGG-**m**IDH1^**high vs low**^ from the Chinese Glioma Genome Atlas (http://www.cgga.org.cn/). Similar to the TCGA analysis, LGG-**m**IDH1^**high**^ from CGGA were enriched in GO related to cell cycle, DNA damage response, and RNA processing (supplement 7). These data elucidate the molecular differences in the high tumor mutational burden within LGG-**m**IDH1 tumors and suggest that GOs belonging to cell cycle regulation and RNA processing may be negatively correlated with mIDH1 patient survival.

**Figure 5.**
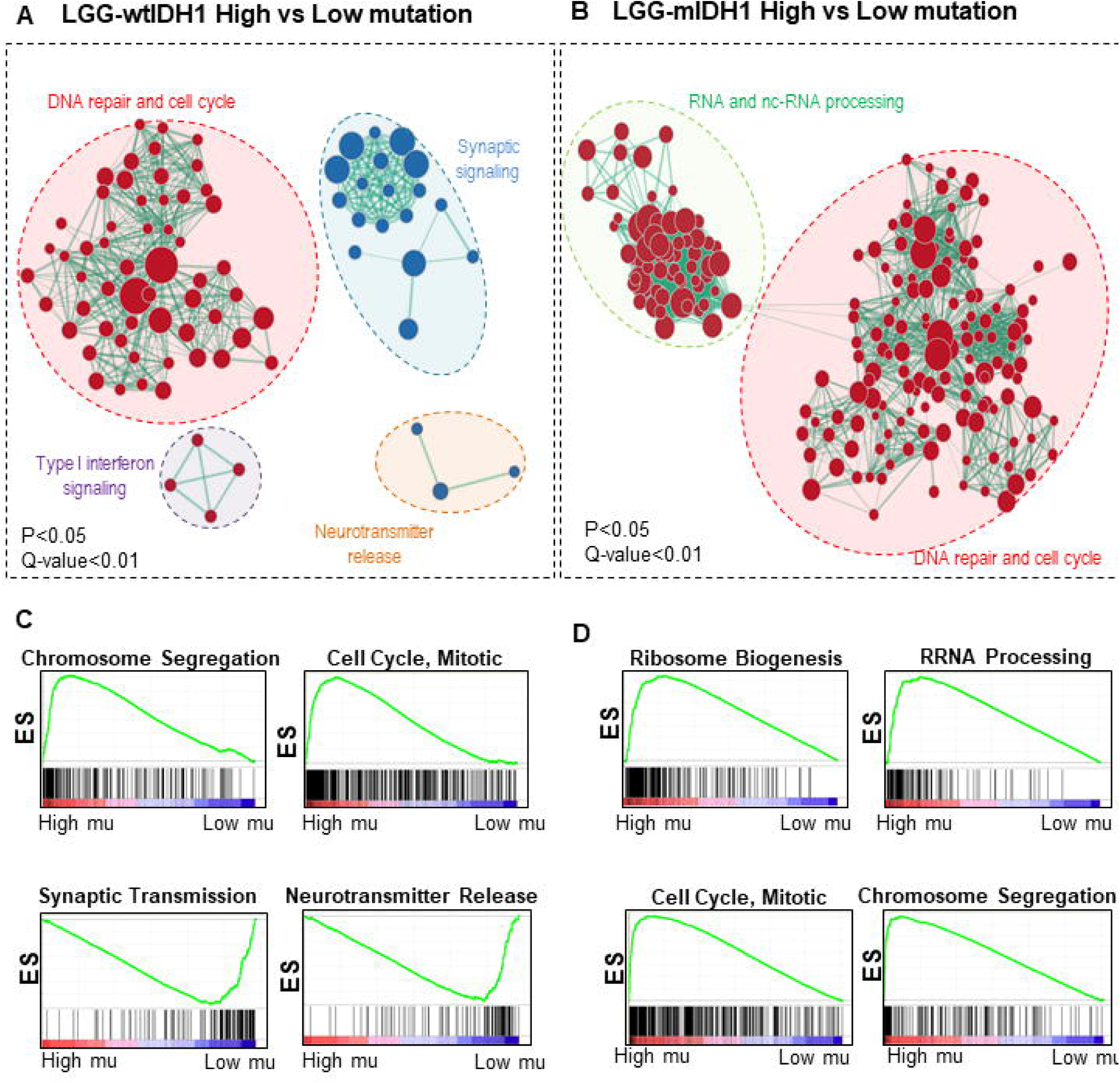
Gene set enrichment analysis (GSEA) of high vs low mutation load in LGG-wtIDH1 and LGG-mIDH1. **(A, B)** Cytoscape map visualization of the positive (red) and negative (blue) enriched GO groups in high vs low mutation load in LGG-**wt**IDH1 **(A)** and LGG-**m**IDH1 **(B). (C, D)** Enrichment plots of the top significantly altered GO in the high vs low mutation load in LGG-**wt**IDH1 and LGG-**m**IDH1.

### Identification of a set of genes that predict survival in LGG-mIDH1 patients

We aimed to construct an expression gene set that could be used to predict survival in LGG-**m**IDH1. To do so, we first eliminated ontologies that were enriched in both the LGG-**wt**IDH1^**high vs. low**^ and LGG-**m**IDH1^**high vs. low**^. The resulting 38 gene sets reflect gene sets selectively enriched in LGG-**m**IDH1^**high**^. LGG-**m**IDH1^**high**^ enriched ontologies were associated with DNA repair, cell cycle-related processes, and chromosomal remodeling. To determine if these gene sets correlate with survival, we used the ‘significance analysis of prognostic signatures’ (SAPS) test (39). SAPS is a powerful tool that computes three p-values (*P*_*pure*_, *P*_*random*_, and *P*_*enrichment*_) for candidate prognostic gene sets, and integrates the three p-values in the form of the SAPS *q*-value. Out of 38 gene sets, only 12 gene sets had a significant SAPS score (*q*-value <0.01). Since these 12 gene sets contain a total of 1483 genes, we performed the ‘least absolute shrinkage and selection operator’ (LASSO) (Fig 6A) to identify those genes within each gene set most associated with survival. From within those 1483 genes, LASSO selected 74 genes that are highly associated with the survival of patients with LGG-**m**IDH1 tumors (Fig 6A). Thus, using TCGA we identify 74 genes within 12 gene sets which predict LGG-**m**IDH1 survival.

**Figure 6.**
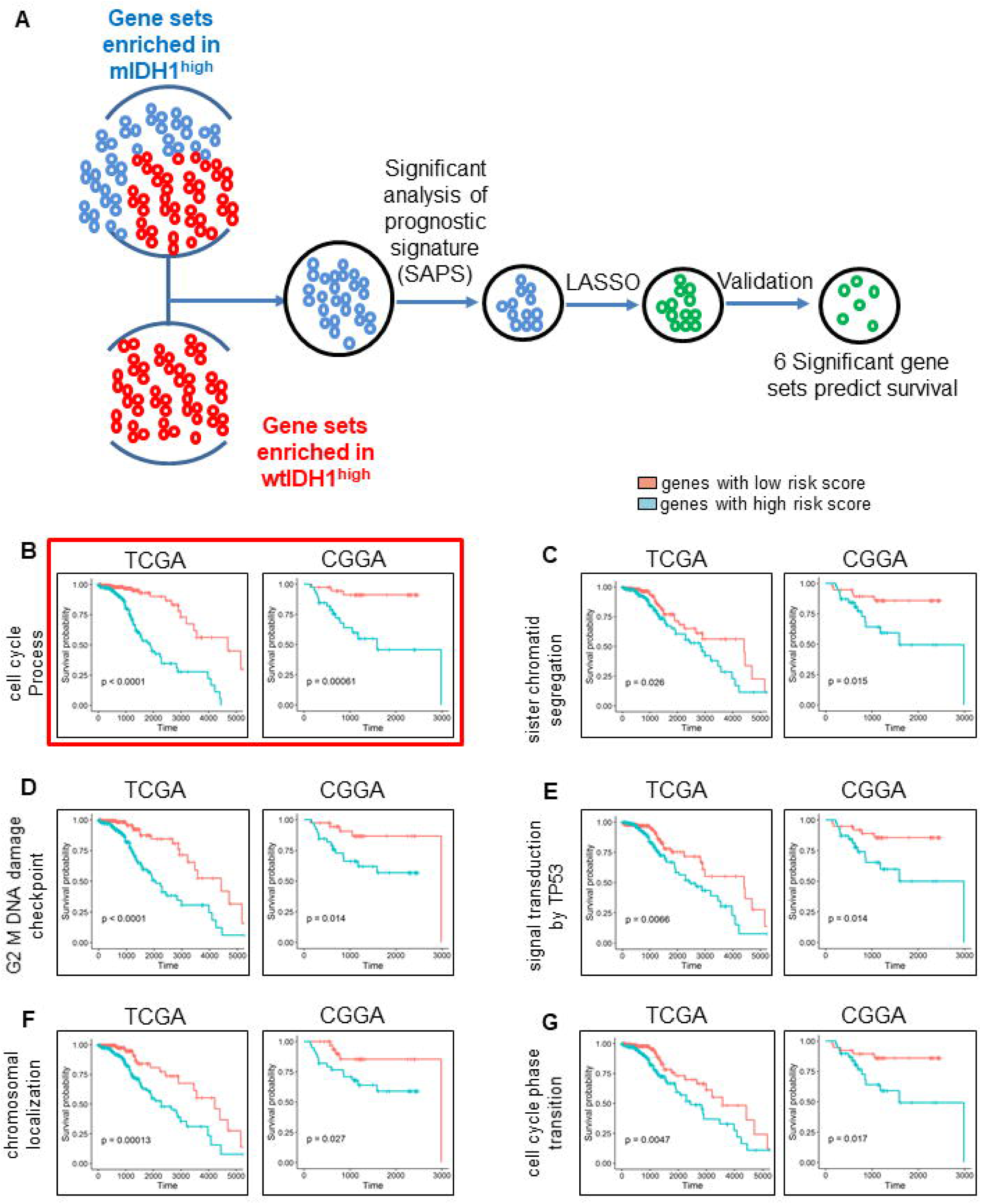
Kaplan-Meier survival of the high risk and low risk genes in LGG-mIDH1. (**A**) Flow chart illustrating the construction of high-risk gene set in LGG-**m**IDH1. Significant gene sets in LGG-**m**IDH1 were selected based om GSEA analysis. SAPS analysis was done to test the significant prognostic gene sets. Finally, LASSO was performed to predict the genes that mostly impact survival. (**B-G**) Kaplan-Meier of the high-risk vs low risk patients based on the 37 candidate genes that predict survival in all 6 gene sets in TCGA (training) and CGGA (validation) dataset. We propose the sequence of 19 genes in **B** as a predictor of survival in LGG-**m**IDH1.

We then validated the twelve gene sets obtained from our TCGA analysis using the CGGA database. LASSO testing of the CGGA database revealed that only six gene sets (37 genes) predicted survival of patients with LGG-**m**IDH1 tumors (Fig 6B-G).

From these six gene sets, we selected the one that was most significant in both databases. This gene set contains 19 genes and predicts survival of patients with LGG-**m**IDH1 tumors with high statistical significance (TCGA: *P*<0.0001, CGGA: *P*<0.00061). The majority of these genes belong to DNA repair pathway (Table 4). Therefore, we propose that this 19 genes’ signature could be utilized as a prognostic marker for survival of patients with LGG-**m**IDH1 tumors.

**Table 4.**
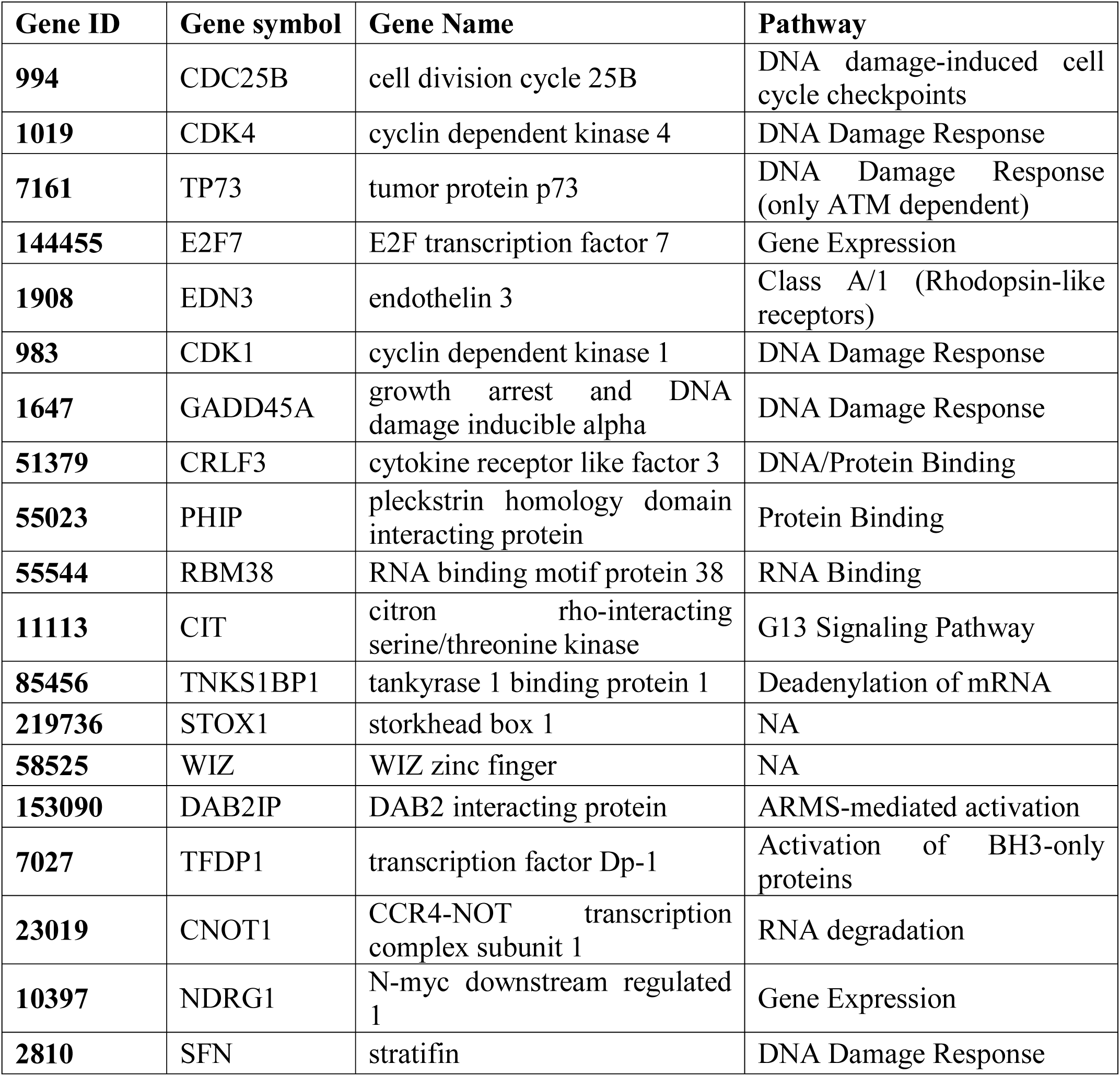
19-genes signature that predicts survival in LGG-mIDH1

| **Gene ID** | **Gene symbol** | **Gene Name** | **Pathway** |
| --- | --- | --- | --- |
| **994** | CDC25B | cell division cycle 25B | DNA damage-induced cell cycle checkpoints |
| **1019** | CDK4 | cyclin dependent kinase 4 | DNA Damage Response |
| **7161** | TP73 | tumor protein p73 | DNA Damage Response (only ATM dependent) |
| **144455** | E2F7 | E2F transcription factor 7 | Gene Expression |
| **1908** | EDN3 | endothelin 3 | Class A/1 (Rhodopsin-like receptors) |
| **983** | CDK1 | cyclin dependent kinase 1 | DNA Damage Response |
| **1647** | GADD45A | growth arrest and DNA damage inducible alpha | DNA Damage Response |
| **51379** | CRLF3 | cytokine receptor like factor 3 | DNA/Protein Binding |
| **55023** | PHIP | pleckstrin homology domain interacting protein | Protein Binding |
| **55544** | RBM38 | RNA binding motif protein 38 | RNA Binding |
| **11113** | CIT | citron rho-interacting serine/threonine kinase | G13 Signaling Pathway |
| **85456** | TNKS1BP1 | tankyrase 1 binding protein 1 | Deadenylation of mRNA |
| **219736** | STOX1 | storkhead box 1 | NA |
| **58525** | WIZ | WIZ zinc finger | NA |
| **153090** | DAB2IP | DAB2 interacting protein | ARMS-mediated activation |
| **7027** | TFDP1 | transcription factor Dp-1 | Activation of BH3-only proteins |
| **23019** | CNOT1 | CCR4-NOT transcription complex subunit 1 | RNA degradation |
| **10397** | NDRG1 | N-myc downstream regulated 1 | Gene Expression |
| **2810** | SFN | stratifin | DNA Damage Response |

## DISCUSSION

Expression of mIDH1 in LGG results in a hypermethylation phenotype and enhanced patients’ survival. In this study, we analyzed the tumor mutational burden, CNVs, mRNA expression, and clinical outcomes utilizing “The Cancer Genome Atlas (TCGA)” database. We validated the results using the Chinese Glioma Genome Atlas (CGGA) database. We propose that tumor mutational burden is a predictor of overall survival in patients with LGG-**m**IDH1 tumors. Further, based on gene expression levels, we constructed a list of “high-risk” genes which we propose could be used as a prognostic marker of overall survival.

Although the total number of mutations in GBM and LGG-**wt**IDH1 tumors is higher than in LGG-**m**IDH1 tumor, the correlation of tumor mutational burden with patient outcome was specific to LGG-**m**IDH1. This suggests that genomic stability is an important contributor to survival in LGG-**m**IDH1 patients, albeit, the role of IDH1^R132H^ on mutation rate in LGG tumors remains to be determined.

Tumor mutational burden has been associated with better response to immunotherapy in many, but not all, tumor types (40). However, the tumor mutational burden can also give rise to intra-tumor heterogeneity which increases treatment resistance, including resistance to immune therapy (41). As recurrent tumors contain an increased tumor mutational burden, and are also more resistant to treatment, and more aggressive than primary tumor (2, 23, 24), there exists a general correlation between tumor mutational burden and tumor progression and aggressiveness.

Previous studies have shown that mutation in PIK3CA and deletion of CDKN2A are associated with poor clinical outcome (30, 31) in LGG. We found that CDKN2A is only present in LGG-**wt**IDH1 (less than 1% of LGG-**m**IDH1), and it was associated with poor clinical outcome in both LGG-**wt**IDH1^**high**^ and LGG-**wt**IDH1^**low**^. Mutation in PIK3CA was present in approximately 7% of LGG-**m**IDH1 patient, but it did not drive unfavorable prognosis in LGG-**m**IDH1^**high**^. Moreover, the frequency of these genetic lesions was not different in the high mutation vs. low mutation group.

GSEA suggests that LGG-**m**IDH1^**high**^ tumors have positive enrichments in cell cycle regulation and DNA repair GO groups as compare to LGG-**m**IDH1^**low**^. This is likely to be a result of genomic instability associated with high tumor mutational burden in LGG-mIDH1. These data are consistent with our recent study which showed that mIDH1 tumor is associated with epigenetic overexpression of genes involved in DNA repair pathway such as ATM (32). At this stage the activation of DNA repair mechanisms in tumors with high mutational burden could be a cause of increased mutation, or its consequence. Future experimental studies will need to address this issue. Another GO which was unique in LGG-**m**IDH1^**high**^ vs LGG-**m**IDH1^**low**^ but not in the LGG-**wt**IDH1 was ‘RNA and non-coding RNA processing’ (42-45). The role of changes in RNA processing, whether causal or effect of high levels of mutation, will also need to be evaluated in forthcoming studies.

In summary, we propose that the tumor mutational burden could be used as a potentially highly reliable marker of overall survival in patients with LGG-**m**IDH1 tumors. Furthermore, we identified a sequence of 19 genes whose expression levels correlate significantly with the OS in this group of patients. As our data were established using the TCGA database and validated using the CGGA database, we propose that our results have high clinical statistical significance. We believe our results are of clinical relevance for therapeutic decisions, and the stratification of patients for clinical trials.

## METHODS

### Source of data and analyses

All clinical, RNAseq, CNVs, and mutational TCGA data were downloaded from the broad institute firebrowse (46) (http://firebrowse.org/?cohort=GBMLGG&download_dialog=true#). CGGA data were downloaded from the CGGA website (http://www.cgga.org.cn/). Patients were categorized according to their mIDH1 status, tumor grade, and tumor mutational load (high vs low). cBioportal platform was used to validate the effect of mutation load on survival of patients with IDH1 mutation. The pan-cancer LGG or GBM cohort were selected and patients were screened for the IDH1 mutation. Classification of mutation load into four groups was done in a web-based analytical tool the cbioportal website. For mutations and CNVs frequency analysis, data from all patients were converted into matrix files and an in-house developed R-script was used to determine the frequency of each mutation in all the analyzed groups.

### Mutation analysis

Mutation analysis was done through the Broad’s institute firebrowse stringent filtering and annotation pipeline to obtain a uniform set of mutation calls (46-49). In brief, the number of mutations and the number of covered bases for each gene were tabulated. The significant metric was calculated for each gene, using the lawrence et al methods (MutSigCV) which measure the significance of mutation burden (50). MutSigCV determines the P value for observing the given quantity of non-silent mutations in the gene, given the background model determined by silent (and noncoding) mutations in the same gene and the neighbouring genes of covariate space (49).

### Survival analysis

Survival data was censored at the last date the patient was known to be alive. Survival functions were estimated by the Kaplan-Meier method and compared using the log-rank test. The median survival time is calculated as the smallest survival time for which the survivor function is less than or equal to 0.5. Cox proportional hazards regression was used to assess the effect of mutation numbers and IDH mutation status on patient survival. In the Cox model, an interaction between the tumor mutational burden and IDH mutation is tested to estimate the effect of mutation numbers with and without the IDH mutation. The assumptions of proportional hazard and linear form of covariates were assessed by martingale residuals plots and the Kolmogorov-type supremum test. All analyses for survival data were done using SAS 9.4 software. P < 0.05 was considered significant.

For regression analysis, only the deceased patients from each group were considered. Linear regression analysis was performed for the “days to death” as dependent variable vs the number of mutated genes per patient (independent variable). Analysis was done using STATA 15.1 software. P < 0.05 was considered significant.

For Significance Analysis of Prognostic Signatures (SAPS), significant gene sets enriched in LGG-**m**IDH1^**high**^ were used to compute the true gene set that can predict survival. SAPS was ran using the Bioconductor R package “SAPS” https://rdrr.io/bioc/saps/man/saps.html.. All gene sets were compared in their ability to predict the survival based on the three P-values computed by SAPS (*P*_*pure*_, *P*_*random*_, and *P*_*enrichment*_) (39). Only gene sets with integrated Q-value <0.01 were considered significant.

### GSEA analysis

All patients were screened for the tumor grade, mutations load, and IDH1 status. Rank file was created by implementing R script from Bader lab (https://github.com/BaderLab/EM-tutorials-docker/blob/master/R_scripts/supplemental_protocol2_rnaseq.R). The gmt file downloaded from broad institute website (http://software.broadinstitute.org/gsea/index.jsp) and it contains all gene ontology (GO) sets to be included In the GSEA analysis. File for positively and negatively enriched groups were used as input files to create the enrichment map in cytoscape.

## Data Availability

Data used were from publicly available databases TCGA, and CGGA.

## ACKNOWLEDGEMENTS

We thank the consulting for statistics, computing and analytics research (CSCAR) at the university of Michigan for the helpful input. We thank the support and academic leadership of Dr. Karin Muraszko, and the administrative and technical support of Angela Collada, and Marta Edwards, respectively. The content is solely the responsibility of the authors.

This work was supported by ‘National Institutes of Health/National Institute of Neurological Disorders & Stroke (NIH/NINDS) Grants [R37-NS094804, R01-NS105556, R21-NS107894 and Rogel Cancer Center Scholar Award to M.G.C.]; NIH/NINDS Grants [R01-NS076991, R01-NS082311, and R01-NS096756 to P.R.L.]; NIH/NIBIB [R01-EB022563 and NCI/UO1-CA-224160 to M.G.C. and P.R.L.]; the Department of Neurosurgery; Leah’s Happy Hearts Foundation, ChadThough Foundation, Pediatric Brain Tumor Foundation, and Smiles for Sophie Forever Foundation [to M.G.C. and P.R.L]. RNA Biomedicine Grant [F046166 to M.G.C.] NIH/NCI [T32-CA009676 Post-doctoral Fellowship to M.S.A].

## ABBREVIATIONS

IDH1: Isocitrate Dehydrogenase1
MS: Median Survival
LGG-mIDH1^high^: Low Grade glioma patients with IDH1 mutation and high mutational burden
LGG-mIDH1^low^: Low Grade glioma patients with IDH1 mutation and low mutational burden
OS: Overall Survival
2HG: 2-Hydroxy glutarate
αKG: α-ketoglutarate
VEGF: Vascular endothelial growth factor
HIF-1α: hypoxia-inducible factor-1α
PIK3CA: Phosphatidylinositol-4,5-Bisphosphate 3-Kinase Catalytic Subunit Alpha
CDKN2A: Cyclin Dependent Kinase Inhibitor 2A
LGG: Low Grade Glioma
HGG: High Grade Glioma
GBM: Glioblastoma Multiforme
TCGA: The Cancer Genome Atlas
CGGA: Chinese Glioma Genome Atlas
Oligo: oligodendrogliomas
Astro: astrocytoma

